# Effectiveness of the 2023 Autumn XBB.1.5 COVID-19 Booster During Summer 2024 in the EU/EEA: A VEBIS Electronic Health Record Network Study

**DOI:** 10.1101/2025.04.30.25326709

**Authors:** James Humphreys, Nathalie Nicolay, Toon Braeye, Izaak Van Evercooren, Christian Holm Hansen, Ida Rask Moustsen-Helms, Chiara Sacco, Massimo Fabiani, Jesús Castilla, Iván Martínez-Baz, Ausenda Machado, Patricia Soares, Rickard Ljung, Nicklas Pihlström, Esther Kissling, Anthony Nardone, Susana Monge, Sabrina Bacci, Baltazar Nunes, VEBIS-EHR working group

## Abstract

We estimated the vaccine effectiveness (VE) of the XBB.1.5 dose given in the autumn 2023 against COVID-19-related hospitalisations and deaths in individuals 65 years of age or older across six EU countries reported following the 2024 summer peak in SARS-CoV-2 test positivity. A historical cohort study was performed by linking electronic health record databases. VE was estimated using Cox regression. Among individuals 65-79 years-old and ≥80 years-old, respectively, VE of the XBB.1.5 dose after ≥6 months post-administration was 13% (95%CI: −12%; 33%) and 7% (95%CI: −7%; 19%) against hospitalisation; and 39% (95%CI: −7%; 65%) and 3% (95%CI: −23%; 23%) against deaths. The 2023 autumnal dose showed very low to no effectiveness at the time of the period of increased SARS-CoV-2 spanning summer 2024.

## Introduction

After the official end of the COVID-19 pandemic, SARS-CoV-2 continues to circulate year-round due to shifting predominance of lineages and sublineages (1). In Europe, following a period of very low activity after the winter of 2023-24, SARS-CoV-2 test positivity began to increase in May 2024. Between May and June 2024, the KP sublineages of JN.1 became predominant in the European Union (EU) (2,3). Although the SARS-CoV-2 KP.2 and KP.3 sublineages were not expected to increase severity (2), concerns focused on their potential for immune escape compared with previously circulating XBB.1.5-like lineages (4–6).

In most EU countries in there was no recommendation for an additional COVID-19 dose in spring 2024. Given waning vaccine effectiveness (VE) and the potential for immune escape, the mismatch in the vaccine component received during the 2023 autumn campaigns and sublineages circulating in spring (BA.2.86/JN.1), protection conferred by the monovalent XBB.1.5 dose was expected to be low in the subsequent months (5,6,8).

A surge in hospitalizations, ICU admissions, and deaths related to laboratory-confirmed COVID-19 followed, with large proportions of these severe cases occurring among individuals aged 65 years and older (1). The exact timing of the peak in COVID-19 deaths and admissions spanning summer 2024 varied by country, though it generally began about eight to nine months after the start of the 2023 autumn vaccination campaigns, during which the monovalent XBB.1.5 dose was most frequently administered (7).

Our objective was to estimate VE of the autumn XBB.1.5 dose against COVID-19-related hospitalisations and deaths occurring between June and August 2024, among individuals eligible for vaccination, aged 65 years or older, and residing in six EU countries.

## Methods

This VE study was undertaken within the VEBIS-EHR network (Vaccine Effectiveness Burden and Impact Studies using Electronic Health Records), a multi-country study funded by the European Centre of Disease Prevention and Control (ECDC) with six participating in the current analysis: Belgium, Denmark, Italy, Spain (Navarre), Portugal, and Sweden.

Detailed methods are available from the study protocol for the 2023-24 season and previous publications (8–10). Briefly, using a common protocol, historical cohorts were constructed by each site, using deterministic linkage of electronic health records. We included individuals aged 65 years or older and eligible to receive the 2023 autumnal COVID-19 dose according to site recommendations. VE was estimated for the period between 1 June to 25 August 2024 using data extracted in October 2024 (9,11).

Individuals eligible for vaccination (Supplementary materials) contributed to unvaccinated person-time at risk if they had not been vaccinated with the 2023 autumn booster between the start and end of the vaccine campaign, as defined locally. Vaccinated individuals contributed to vaccinated person-time at risk after 14 days post-vaccination had elapsed. Individuals’ person-time at risk stopped at the date of the outcome, date of death for any cause, on the date of a subsequent vaccine dose during the autumn campaign, or at the end of the study period (25 August 2024).

Hospitalisation due to COVID-19 was defined as a hospital admission due to a severe acute respiratory infection with a SARS-CoV-2 positive test from 14 days before to 1 day after admission or with COVID-19 as the main diagnosis in hospital admission or discharge records. A COVID-19-related death was defined as death with main cause coded as COVID-19, and/or with a SARS-CoV-2 positive laboratory result in the 30 days preceding death.

Each study site used proportional hazards Cox regression models, with vaccination status as time-varying exposure and calendar time as the underlying scale, to estimate vaccine hazard ratios (aHR) adjusted by 5-year age-group, sex, comorbidities, and previous number of vaccine booster doses, with other covariates included according to study site-specific protocols. We pooled study site-specific hazard ratios using a random-effects meta-analysis. We estimated pooled VE as one minus the pooled hazard ratio estimates expressed as a percentage (VE=(1-pooled aHR)*100), with heterogeneity scored using the I^2^ index. Analysis was stratified by age group (65-79 years old and 80 and older)

## Results

We included 19.3 million participants in this analysis. At the end of the study period, 13.3 million participants were unvaccinated and 6.0 million participants had received the XBB.1.5 vaccine during autumn 2023. Among those vaccinated, 99.8% of the participants had received the vaccine more than 6 months earlier (Table 1). Compared with unvaccinated participants, those who had received the monovalent XBB.1.5 vaccine presented a higher prevalence of high-risk comorbidities (6.6% vs 2.1%) and a higher proportion of those with at least two booster doses before the start of the autumn 2023 campaign (92.7% vs 32.5%).

**Table 1.** Descriptive characteristics of the study population (N= 19,306,009) by vaccination status and time since vaccination at the end of the study period*, within the six study sites (Belgium, Denmark, Italy, Navarre-Spain, Portugal, and Sweden), from 1 June 2024 to 25 August 2024, VEBIS-EHR network.

| Variable | Not vaccinated<br>n (%) | Vaccinated ≥14 days<br>n (%) | Vaccinated 14-89<br>days<br>n (%) | Vaccinated 90-179 days<br>n (%) | Vaccinated ≥180<br>days<br>n (%) |
| --- | --- | --- | --- | --- | --- |
| Total (row % over<br>total = 19,306,009) | 13,264,417 (68.7) | 6,041,592 (31.3) | 67 (0.0) | 13,455 (0.1) | 6,028,070 (31.2) |
| Study site |  |  |  |  |  |
| Belgium | 1,009,248 (7.6) | 1,091,923 (18.1) | 0 | 0 (0.0) | 1,091,923 (18.1) |
| Denmark | 211,557 (1.6) | 878,421 (14.5) | 0 | 77 (0.6) | 878,344 (14.6) |
| Italy | 10,580,504 (79.8) | 1,279,075 (21.2) | 0 | 4,649 (34.6) | 1,274,426 (21.1) |
| Navarre | 44,974 (0.3) | 81,247 (1.3) | 0 | 655 (4.9) | 80,592 (1.3) |
| Portugal | 936,703 (7.1) | 1,269,745 (21.0) | 67 (100) | 4,981 (37) | 1,264,697 (21.0) |
| Sweden | 481,431 (3.6) | 1,441,181 (23.9) | 0 | 3,093(23) | 1,438,088 (23.9) |
| Age group (years) |  |  |  |  |  |
| 65-79 | 9,434,356 (71.1) | 4,130,612 (68.4) | 44 (65.7) | 8,951 (66.5) | 4,121,617 (68.4) |
| ≥80 | 3,830,061 (28.9) | 1,910,980 (31.6) | 23 (34.3) | 4,504 (33.5) | 1,906,453 (31.6) |
| Sex |  |  |  |  |  |
| Male | 7,417,948 (55.9) | 3,203,628 (53.0) | 37 (55.2) | 7,049 (52.4) | 3,196,542 (53.0) |
| Female | 5,846,443 (44.1) | 2,837,362 (47.0) | 30 (44.8) | 5,829 (43.3) | 2,831,503 (47.0) |
| Missing | 26 (0.0) | 602 (0.0) | 0 (0.0) | 577 (4.3) | 25 (0.0) |
| Comorbidities** |  |  |  |  |  |
| High risks | 280,976 (2.1) | 400,625 (6.6) | 18 (26.9) | 1,292 (9.6) | 399,315 (6.6) |
| Medium/low risk | 4,702,692 (35.5) | 3,058,940 (50.6) | 30 (44.8) | 6,317 (46.9) | 3,052,593 (50.6) |
| No | 8,251,984 (62.2) | 2,565,887 (42.5) | 15 (22.4) | 5,266 (39.1) | 2,560,606 (42.5) |
| Missing | 28,765 (0.2) | 16,140 (0.3) | <5 (6.0) | 580 (4.3) | 15,556 (0.3) |
| Number of<br>previous booster<br>doses |  |  |  |  |  |
| 0 | 1,403,530 (10.6) | 29,279 (0.5) | 0 (0.0) | 196 (1.5) | 29,083 (0.5) |
| 1 | 7,546,294 (56.9) | 411,648 (6.8) | 10 (14.9) | 1,724 (12.8) | 409,914 (6.8) |
| 2 | 3,898,637 (29.4) | 3,831,236 (63.4) | 41 (61.2) | 7,489 (55.7) | 3,823,706 (63.4) |
| 3 | 397,923 (3.0) | 1,420,043 (23.5) | 11 (16.4) | 3,107 (23.1) | 1,416,925 (23.5) |
| 4 | 17,983 (0.1) | 347,173 (5.7) | 0 (0.0) | 358 (2.7) | 346,815 (5.8) |
| 5 | 46 (0.0) | 1,599 (0.0) | 0 (0.0) | 0 (0.0) | 1,599 (0.0) |
| Missing | <5 (0.0) | 614 (0.0) | 5 (7.4) | 581 (4.3) | 28 (0.0) |
| Vaccine product |  |  |  |  |  |
| Pfizer (XBB.1.5) | - | 5,942,540 (98.4) | 67 (100%) | 12,650 (94.0) | 5,929,823 (98.4) |
| Moderna (XBB.1.5) | - | 49,089 (0.8) | 0 (0.0) | 0 (0.0) | 49,089 (0.8) |
| Novavax (XBB.1.5) | - | 5,165 (0.1) | 0 (0.0) | 220 (1.6) | 4,945 (0.1) |
| Others | - | 44,177 (0.7) | 0 (0.0) | 0 (0.0) | 44,177 (0.7) |
| Missing | - | 621 (0.0) | 0 (0.0) | 585 (4.3) | 36 (0.0) |

\* Vaccination status and time since vaccination were assessed at the end of the individual observation period
\*\* High risk comorbidities: immunocompromised conditions with COVID-19 vaccine recommendation; Medium/low risk: non-immunocompromised conditions with COVID-19 vaccine recommendation; No comorbidities: persons without any of the risk comorbidities. (Details are presented in Table S3).

Among participants aged 65–79 years, the unvaccinated and vaccinated groups had COVID-19 hospitalisation rates of 6.2 per 100,000 person-months (1,664 hospitalisations over 26.7 million person-months) and 6.4 per 100,000 person-months (748/11.7 million person-months), respectively. Corresponding COVID-19-related death rates were 0.9 per 100,000 person-months (224/24.4 million person-months) and 2.1 per 100,000 person-months (191/9.3 million person-months), respectively. Overall, XBB.1.5 vaccine effectiveness (VE) against COVID-19 hospitalisations was 13% (95% CI: –12% to 33%). Stratified by time since vaccination, VE was 16% (95% CI: –52% to 54%) among those 90–179 days post-vaccination and 13% (95% CI: –12% to 33%) among those ≥180 days post-vaccination. Due to the low number of events, VE could not be estimated for the 14–89-day interval. Against COVID-19-related deaths, VE was 38% (95% CI: –12% to 65%) for participants vaccinated ≥180 days ago. No other VE estimates could be produced by time since vaccination for the 65–79-year age group (Table 2).

**Table 2.**
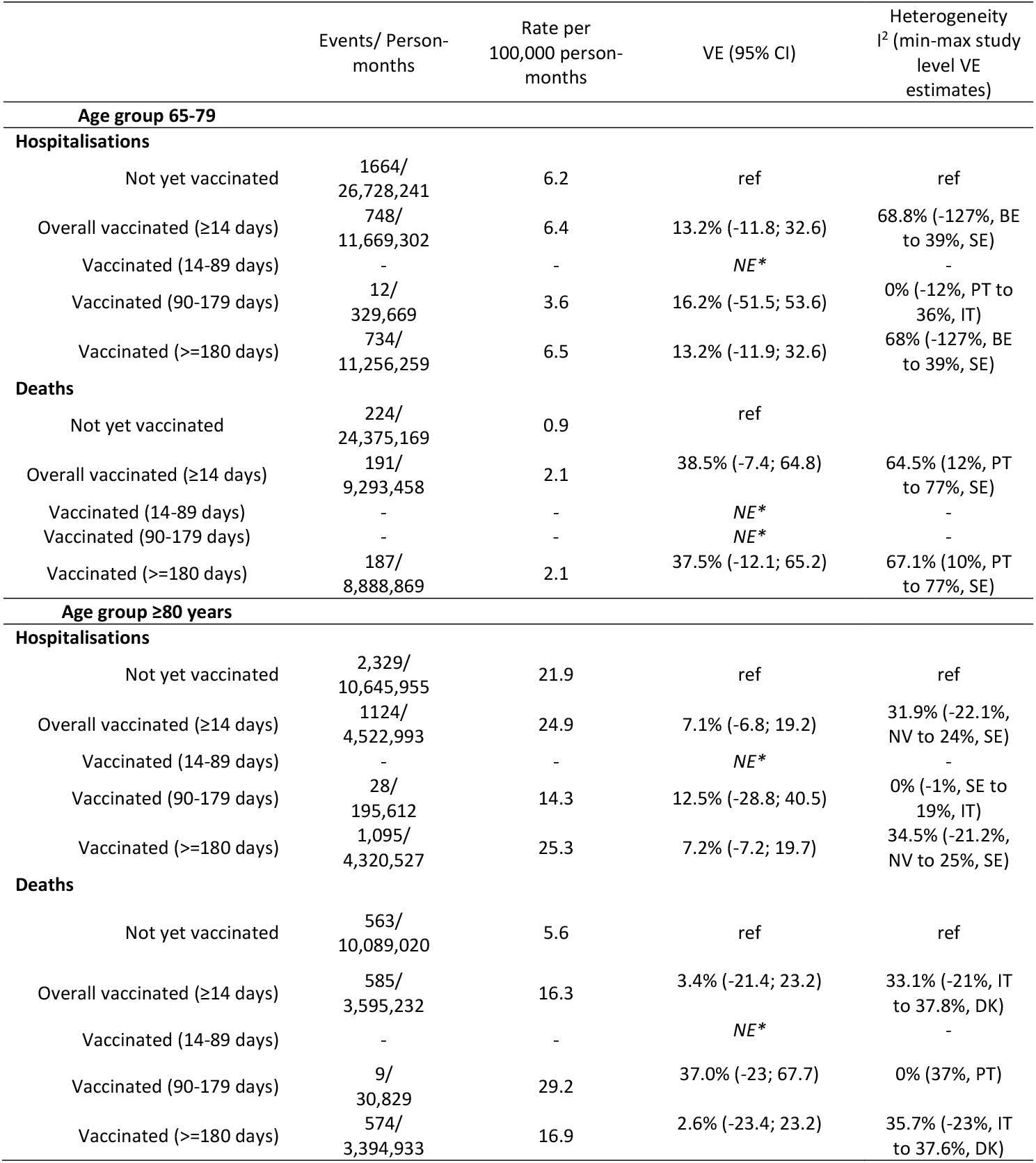
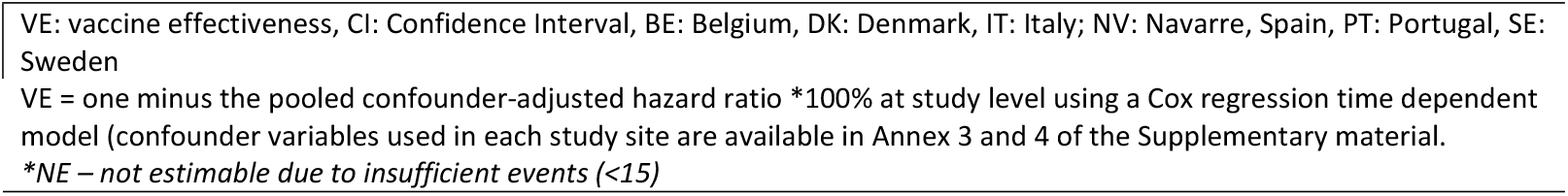
Number of COVID-19 hospitalisations and COVID-19-related deaths, person months at risk by vaccine status, and VE overall and by time since vaccination for individuals aged 65-79 and ≥80 years old, within the six study sites (Belgium, Denmark, Italy, Navarre-Spain, Portugal, and Sweden), from 1 June 2024 to 25 August 2024, VEBIS-EHR network

|  | Events/ Person-months | Rate per<br>100,000 person-months | VE (95% CI) | Heterogeneity<br>I<sup>2</sup> (min-max study<br>level VE<br>estimates) |
| --- | --- | --- | --- | --- |
| **Age group 65-79** |  |  |  |  |
| **Hospitalisations** |  |  |  |  |
| Not yet vaccinated | 1664/<br>26,728,241 | 6.2 | ref | ref |
| Overall vaccinated (≥14 days) | 748/<br>11,669,302 | 6.4 | 13.2% (-11.8; 32.6) | 68.8% (-127%, BE<br>to 39%, SE) |
| Vaccinated (14-89 days) | - | - | NE* | - |
| Vaccinated (90-179 days) | 12/<br>329,669 | 3.6 | 16.2% (-51.5; 53.6) | 0% (-12%, PT to<br>36%, IT) |
| Vaccinated (≥180 days) | 734/<br>11,256,259 | 6.5 | 13.2% (-11.9; 32.6) | 68% (-127%, BE<br>to 39%, SE) |
| **Deaths** |  |  |  |  |
| Not yet vaccinated | 224/<br>24,375,169 | 0.9 | ref |  |
| Overall vaccinated (≥14 days) | 191/<br>9,293,458 | 2.1 | 38.5% (-7.4; 64.8) | 64.5% (12%, PT<br>to 77%, SE) |
| Vaccinated (14-89 days) | - | - | NE* | - |
| Vaccinated (90-179 days) | - | - | NE* | - |
| Vaccinated (≥180 days) | 187/<br>8,888,869 | 2.1 | 37.5% (-12.1; 65.2) | 67.1% (10%, PT<br>to 77%, SE) |
| **Age group ≥80 years** |  |  |  |  |
| **Hospitalisations** |  |  |  |  |
| Not yet vaccinated | 2,329/<br>10,645,955 | 21.9 | ref | ref |
| Overall vaccinated (≥14 days) | 1124/<br>4,522,993 | 24.9 | 7.1% (-6.8; 19.2) | 31.9% (-22.1%,<br>NV to 24%, SE) |
| Vaccinated (14-89 days) | - | - | NE* | - |
| Vaccinated (90-179 days) | 28/<br>195,612 | 14.3 | 12.5% (-28.8; 40.5) | 0% (-1%, SE to<br>19%, IT) |
| Vaccinated (≥180 days) | 1,095/<br>4,320,527 | 25.3 | 7.2% (-7.2; 19.7) | 34.5% (-21.2%,<br>NV to 25%, SE) |
| **Deaths** |  |  |  |  |
| Not yet vaccinated | 563/<br>10,089,020 | 5.6 | ref | ref |
| Overall vaccinated (≥14 days) | 585/<br>3,595,232 | 16.3 | 3.4% (-21.4; 23.2) | 33.1% (-21%, IT<br>to 37.8%, DK) |
| Vaccinated (14-89 days) | - | - | NE* | - |
| Vaccinated (90-179 days) | 9/<br>30,829 | 29.2 | 37.0% (-23; 67.7) | 0% (37%, PT) |
| Vaccinated (≥180 days) | 574/<br>3,394,933 | 16.9 | 2.6% (-23.4; 23.2) | 35.7% (-23%, IT<br>to 37.6%, DK) |

VE: vaccine effectiveness, CI: Confidence Interval, BE: Belgium, DK: Denmark, IT: Italy; NV: Navarre, Spain, PT: Portugal, SE: Sweden
VE = one minus the pooled confounder-adjusted hazard ratio \*100% at study level using a Cox regression time dependent model (confounder variables used in each study site are available in Annex 3 and 4 of the Supplementary material.
*\*NE – not estimable due to insufficient events (<15)*

Among participants aged ≥80 years, COVID-19 hospitalisation rates were 21.9 per 100,000 person-months (2,329/10.6 million person-months) in the unvaccinated group and 24.9 per 100,000 person-months (1,124/4.5 million person-months) in the vaccinated group. COVID-19-related death rates were 5.6 per 100,000 person-months (563/10.1 million person-months) among the unvaccinated and 16.3 per 100,000 person-months (585/3.6 million person-months) in the vaccinated group (Table 2). For those ≥180 days post-vaccination, VE was 7% (95% CI: –7% to 19%) against hospitalisations and 3% (95% CI: –21% to 23%) against deaths. For those vaccinated 90–179 days earlier, VE was 13% (95% CI: –29% to 41%) against hospitalisations and 37% (95% CI: –23% to 68%) against deaths.

Heterogeneity between estimates produced at the study-site level and later pooled via random effects was moderate (I^2: 65%–69%) for the 65–79-year age group and low (I^2: 0%–36%) for those ≥80 years (Table 2).

## Discussion

Within the VEBIS EHR network, we estimated null or very low XBB.1.5 VE among individuals aged ≥65 years between June and August 2024. Nearly all (99.8%) vaccinated participants had received their dose at least six months prior. During the study period, XBB.1.5 VE against COVID-19 hospitalisations was 13% (95% CI: –12% to 33%) among those aged 65–79 years and 7% (95% CI: –7% to 19%) among those ≥80 years. Against COVID-19-related deaths, VE was 39% (95% CI: –7% to 65%) in the 65–79-year age group and 3% (95% CI: –21% to 23%) in those aged ≥80 years.

These findings are consistent with two studies examining XBB.1.5 VE against COVID-19 hospitalisation during summer 2024 in older populations (12,13). In a Canadian study covering ten months following the start of the 2023 vaccination campaign, XBB.1.5 vaccination conferred null or very low (<14%) protection ≥7 months post-vaccination, including during spring/summer 2024 (12). Similarly, the UK Health Security Agency (UKHSA) reported autumn 2023 VE estimates of 32% (95% CI: 11–48%), 25% (95% CI: 15–35%), and 12% (95% CI: 4–20%) for individuals who were 20–24 weeks (140–168 days), 25–29 weeks (175–203 days), and ≥30 weeks (210 days) post-vaccination, respectively, in April 2024 (13).

The lower XBB.1.5 VE observed among older EU residents during summer 2024 may be due to waning protection beyond six months (8,12), as well as antigenic mismatches between the XBB.1.5 vaccine component and the Omicron BA.2.86 sublineages (including JN.1) circulating between June and August 2024 (5,6; Supplementary material). In two previous VEBIS-EHR studies that estimated VE during XBB.1.5 (9) and BA.2.86/JN.1 (10) predominance, we noted decreases in VE against severe outcomes, possibly reflecting both viral evolution and the natural waning of vaccine-induced immunity.

Nonetheless, the XBB.1.5 vaccine continued to provide moderate protection during the BA.2.86 predominance period (10). In England, where XBB.1.5 was offered as a 2024 spring booster dose, protection against hospitalisations in the first two months following vaccination returned to levels observed early in the 2023 autumn campaign, which supports waning immunity (rather than immune escape by emerging KP sublineages) as the primary explanation for declining protection (13).

When interpreting these results, it is important to consider the limitations inherent to using secondary electronic health record (EHR) data for VE research—namely, misclassification of vaccination status and outcomes, and the absence of sufficient covariates for full confounder adjustment. Additionally, because we estimated VE approximately eight months after the 2023 autumn campaign, there is a higher risk of bias from differential depletion of susceptibles by vaccination status (14,15), possibly underestimating VE. This bias could partly explain the unexpected observation of higher COVID-19 hospitalisation and death rates among vaccinated individuals (Table 2). However, simulation studies suggest that this bias alone is unlikely to fully account for the decline in VE with time since vaccination or the elevated incidence among vaccinated persons (16).

There is an additional limitation impacting estimates in one participating country, Belgium, for which the national demographic dataset used to censor deaths in linked registry analyses has not been updated since July 2024. Records after this date may include some individuals who have since died as they can no longer be censored appropriately, potentially leading to an underestimation of VE against hosptialisation. However, Belgium contributed a relatively small proportion of the total events across all participating sites in this analysis, and are not included in estimates against COVID-19 related mortality. Therefore, while this issue must be taken into account when interpreting the results, its overall impact on the multicountry VE estimates is likely to be limited.

In conclusion, our multicountry study indicates that beyond six months after administration, the XBB.1.5 vaccine provides minimal residual protection against severe COVID-19 outcomes among older adults during the summer 2024 peak in COVID-19 activity. These findings highlight the need for improved COVID-19 vaccines offering longer-lasting effectiveness. Should COVID-19 circulation follow a predictable seasonal pattern, the results may support considerations for a spring booster dose in higher-risk groups.

## Ethics

All study sites participating in this study conformed with their respective national and EU ethical and data protection requirements. Ethical statements for each of the participating study sites:

### Belgium

Data linkage and collection within the data-warehouse have been approved by the information security committee. The study was conducted in accordance with the Declaration of Helsinki. Ethical approval was granted for the gathering of data from hospitalised patients by the Committee for Medical Ethics from the Ghent University Hospital (reference number BC-07507) and authorisation for possible individual data linkage using the national register number from the Information Security Committee (ISC) Social Security and Health (reference number IVC/KSZG/20/384). Linkage of hospitalised patient data to vaccination and testing within the LINK-VACC project was approved by the Medical Ethics Committee UZ Brussels–VUB on 3 February 2021 (reference number 2020/523), and authorisation from the ISC Social Security and Health (reference number IVC/KSZG/21/034).

### Denmark

Only administrative register data was used for the study. According to Danish law, ethics approval is exempt for such research, and the Danish Data Protection Agency, which is dedicated ethics and legal oversight body, thus waives ethical approval for the study of administrative register data when no individual contact of participants is necessary, and only aggregate results are included as findings. The study is, therefore, fully compliant with all legal and ethical requirements, and there are no further processes available regarding such studies.

### Navarre (Spain)

The study was approved by Navarre’s Ethical Committee for Clinical Research, which waived the requirement of obtaining informed consent.

### Portugal

The study received approval from the Ethical Committee and the Data Protection Officer of the Instituto Nacional de Saúde Doutor Ricardo Jorge. Given that data was irreversibly anonymised, the need for the participants’ informed consent was waived by the Ethical Committee.

### Italy

This study, based on routinely collected data, will not be submitted for approval to an ethical committee because the dissemination of COVID-19 surveillance data was authorised by the Italian law N. 52 of 19 May 2022, following the law decree N. 24 of 24 March 2022 (Article n. 13). Based on the same acts, the information on COVID-19 vaccination was retrieved by the Italian National Institute of Health using data from the National Immunisation Information System of the Italian Ministry of Health. Because of the retrospective design and the large size of the population under study, in accordance with the Authorisation n. 9 released by the Italian data protection authority on 15 December 2016, the individual informed consent was not requested for the conduction of this study.

### Sweden

The Swedish study is approved by the Swedish Ethical Review Authority (2020-06859, 2021-02186) and has conformed to the principles embodied in the Declaration of Helsinki. Consent to participate is not applicable as this is a register-based study.

## Funding

All the public health organisations involved received funding from the European Centre for Disease Prevention and Control (ECDC) implementing Framework Contract ECDC/2021/018 ‘Vaccine effectiveness and impact of COVID-19 vaccines through routinely collected exposure and outcome using health registries’ (RS/2022/DTS/24104). In Portugal, this work was also supported by FCT – Fundação para a Ciência e Tecnologia, I.P. by project reference CEECINST/00049/2021/CP2817/CT0001 and DOI identifier 10.54499/CEECINST/00049/2021/CP2817/CT0001

## Data Availability

Authors cannot share the data used for this study, which should be requested to the data owner institutions following their respective procedures.

## Conflict of interest

Authors declare no competing interests.

## Author contributions

S Bacci, N Nicolay, J Humphreys, B Nunes, and S Monge conceived the study. J Humphreys, B Nunes, N Nicolay and S Monge conceived the methods. All authors from Public Health institutions at each study site were responsible for data management and analysis at the study site level. J Humphreys was responsible for pooling site level estimates. J Humphreys drafted the manuscript, with the help of B Nunes. All authors contributed to the interpretation of the results and critically reviewed the manuscript. All authors approved the final version of this manuscript. All authors within the VEBIS-EHR working group made a substantial contribution to the conception or design of the work, critically revised the manuscript, provided their final approval of the version to be published, and agreed to be accountable for all aspects of the work.

## Supplementary material Definition 1: Eligibility criteria

The study population includes community-dwelling individuals ≥65 years of age in national EHR databases. The study population should be eligible for seasonal COVID-19 vaccination, including belonging to an age group for whom seasonal COVID-19 vaccination has been recommended in each site/country. Eligibility will be based on the following criteria as of the first day of the vaccination campaign, which may be adapted to match national recommendations:

- Aged between 65 and 110 years at the beginning of the vaccination campaign, or belonging to an age group over 65 years for which the vaccine dose being evaluated has been recommended, if different (it should be recommended for the entire age group). Birth year may be used instead of age in countries where vaccine recommendations are based on birth cohort, or where only year of birth is available.
- Permanent resident in the EU/EEA territory covered in the study (for each study site, according to the most recent information).
- Not residents of a nursing home/long term care facility (according to the most recent information at the beginning of the seasonal vaccination campaign).
- Received their first ever COVID-19 vaccine dose as part of an age-specific vaccination campaign (i.e., excluding those vaccinated before it was generally recommended in the corresponding age-group or, alternatively, excluding the first 5% of persons vaccinated within each age-group – for each 5-year age bracket-as these first vaccinees may not be representative of their corresponding age group).
- Completed primary vaccination at least 180 days before the start of the seasonal vaccination campaign.
- Has not received a COVID-19 vaccine dose, irrespective of the number of doses, in the last 90 days before the start of the seasonal vaccination campaign; has no documented SARS-CoV-2 infection (nor has been hospitalised due to COVID-19) in the 90 days before the start of the seasonal vaccination campaign (25), or; other criteria following the relevant national guidelines of each study site (for example, if seasonal vaccine is recommended irrespective of the time since last infection).
- Does not have inconsistent or missing data on vaccination (vaccination status unknown, any vaccination date is unknown, any vaccine brand is unknown, number of doses is unknown, interval between primary course first and second dose is shorter than 19 days, interval between complete primary vaccination and booster dose or between booster doses is shorter than 90 days, number of doses higher than recommended, received any vaccine brand not approved by EMA, or the combination of vaccine brands is not a recommended schedule -may vary by age group).

**Figures S1-4. Forest plots by age group and outcome, pooled estimates including the six study sites (Belgium, Denmark, Italy, Navarre-Spain, Portugal, and Sweden), from 1 June 2024 to 25 August 2024, VEBIS-EHR network**.

**Figure S1.**
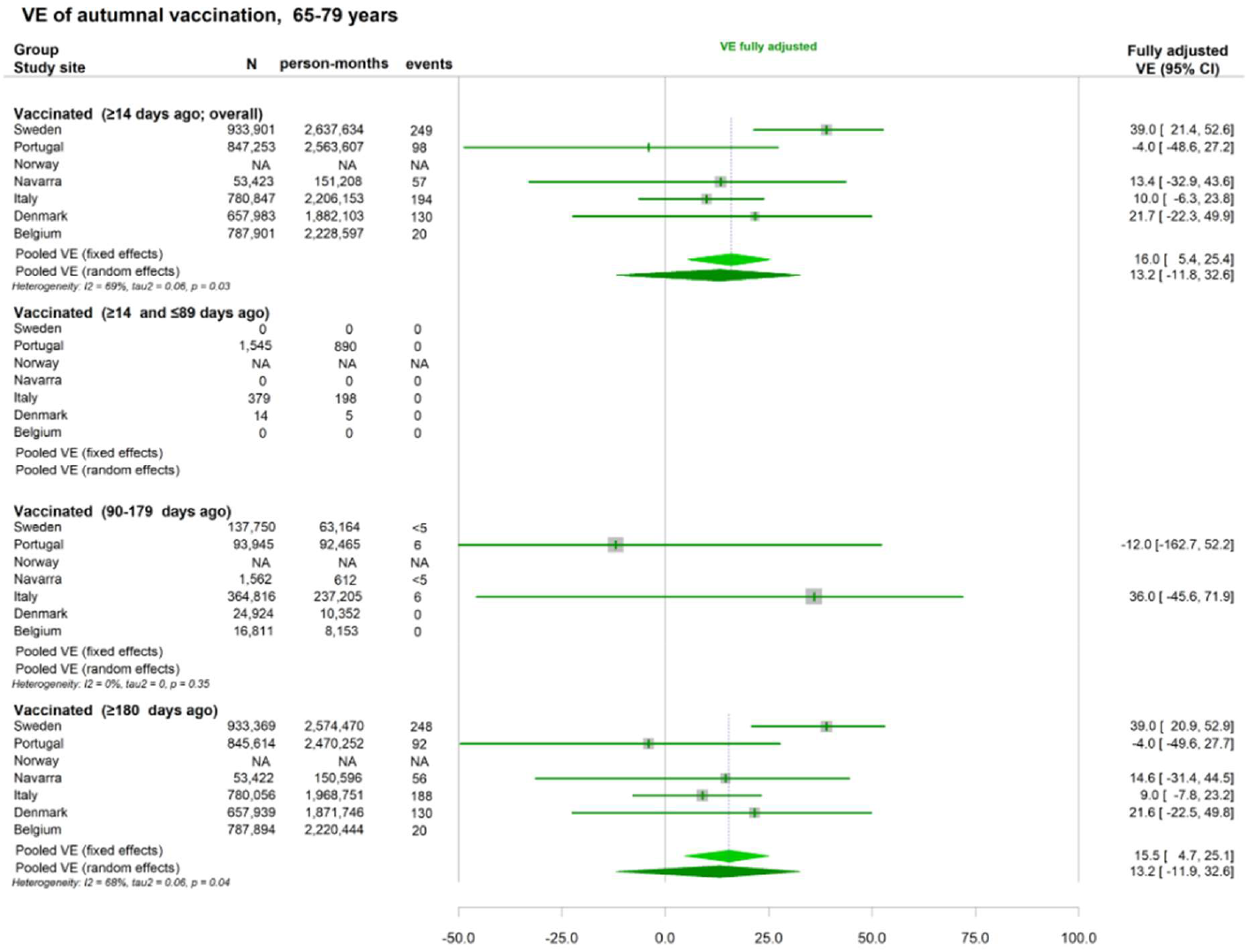
Forest plot of autumn vaccination VE against hospitalisation related to COVID-19 among those aged 65-79 years.

**Figure S2.**
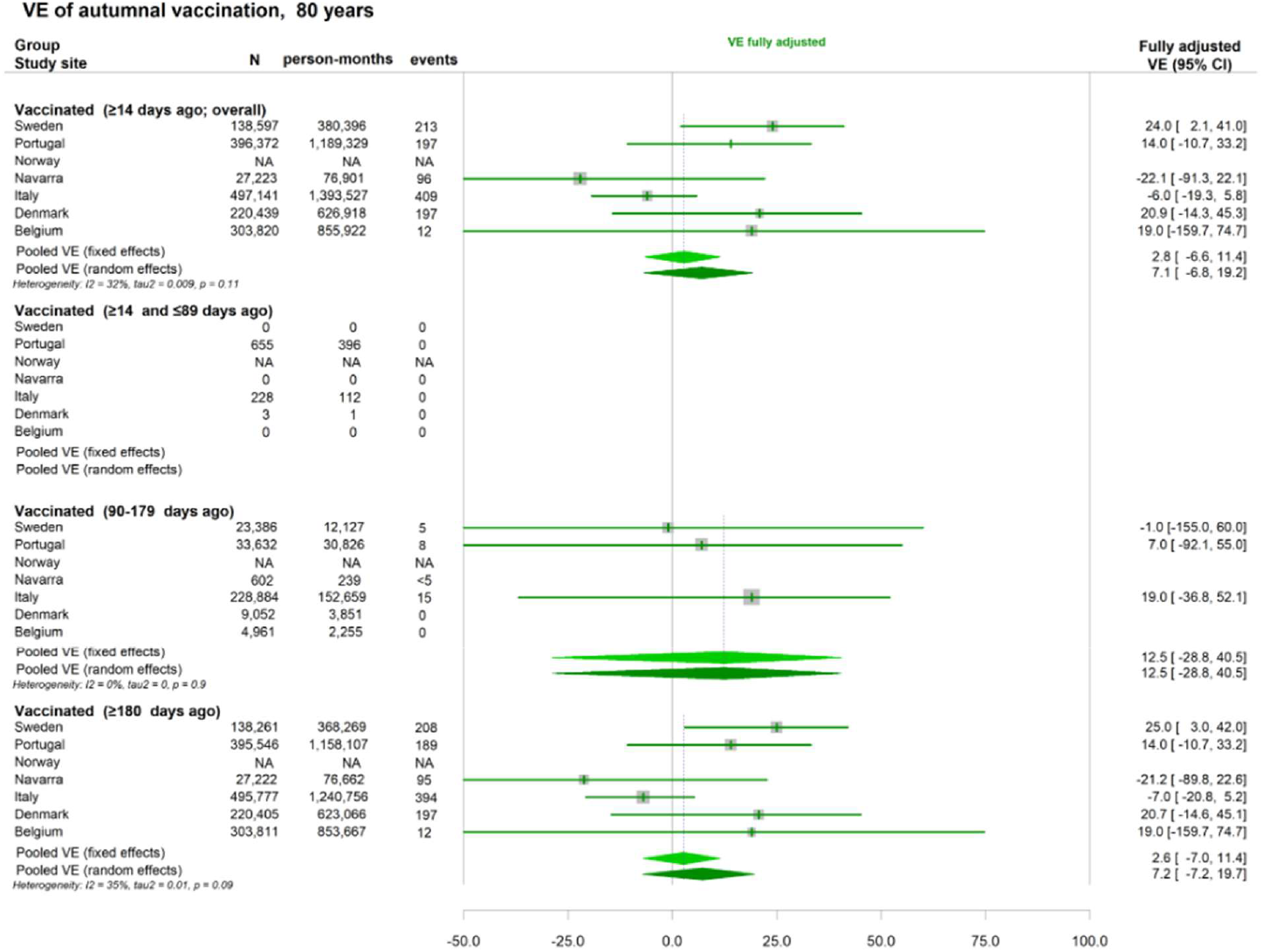
Forest plot of autumn vaccination VE against hospitalisation related to COVID-19 among those aged 80 plus.

**Figure S3.**
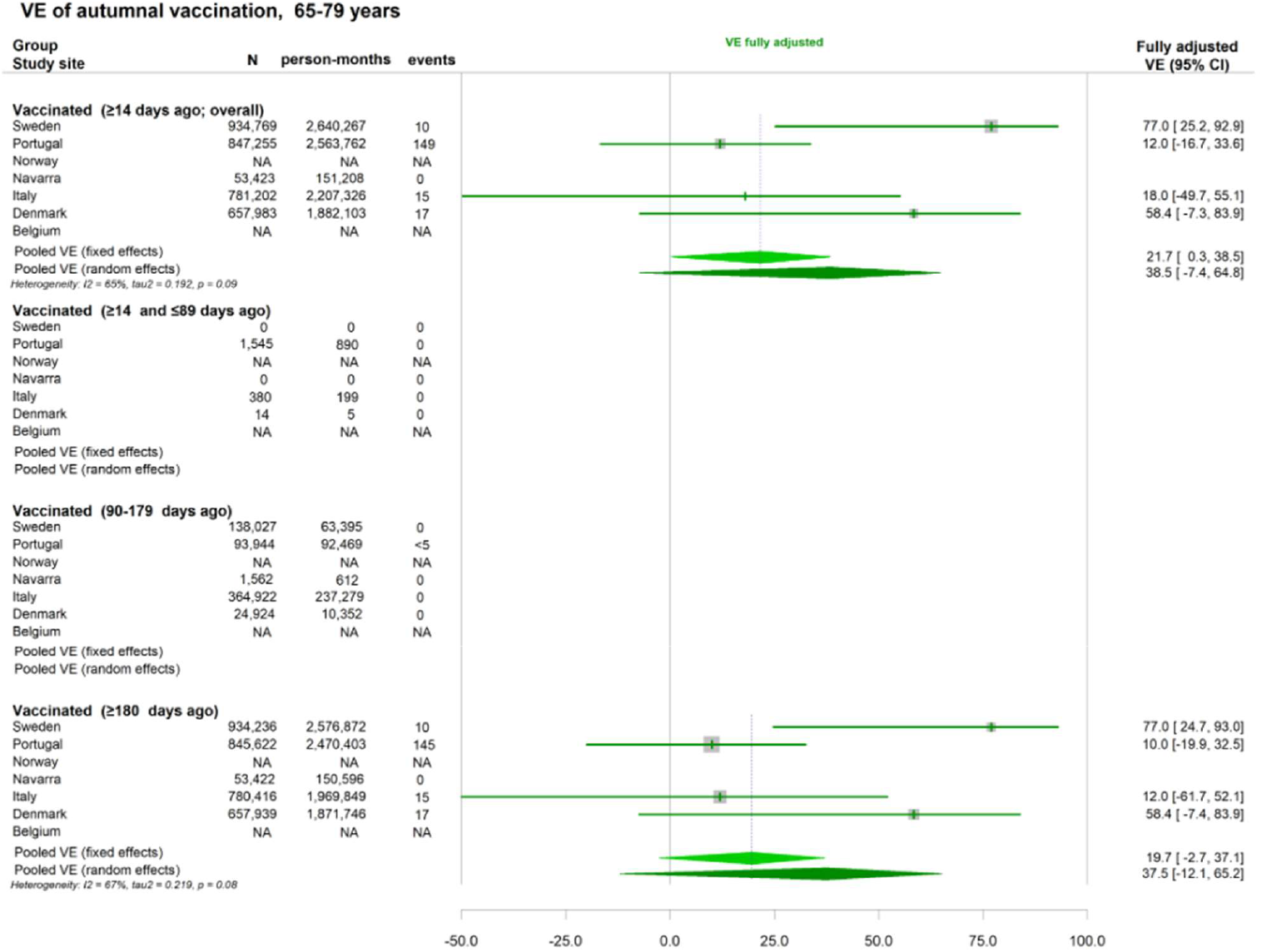
Forest plot of autumn vaccination VE against death due to COVID-19 among those aged 65-79 years.

**Figure S4.**
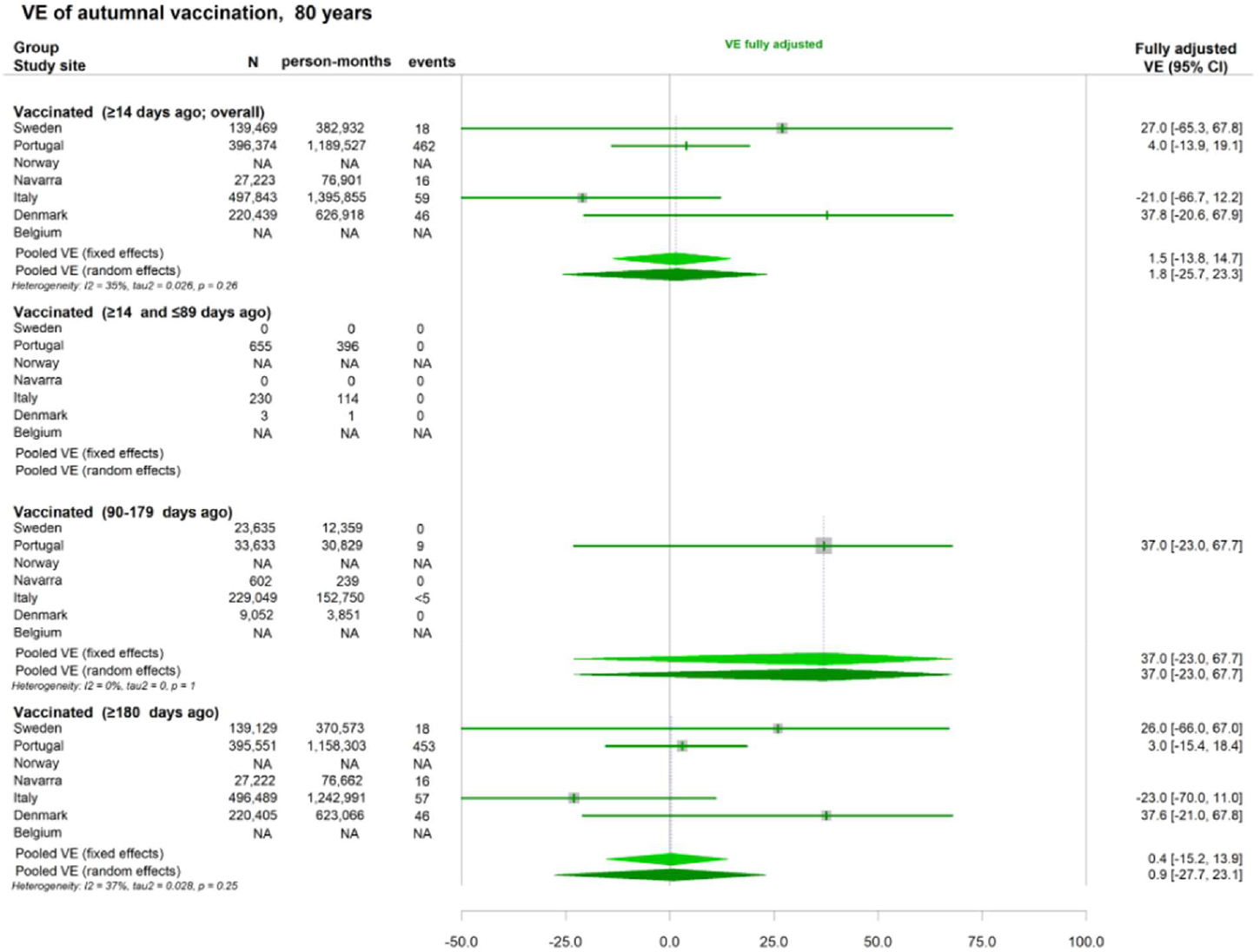
Forest plot of autumn vaccination VE against death due to COVID-19 among those aged 80 plus.

## Notes

### Competing Interest Statement

The authors have declared no competing interest.

### Clinical Protocols

https://www.ecdc.europa.eu/en/publications-data/protocol-covid-19-vaccine-effectiveness-estimation-using-health-data-registries

### Author Declarations

Belgium: Data linkage and collection within the data-warehouse have been approved by the information security committee. The study was conducted in accordance with the Declaration of Helsinki. Ethical approval was granted for the gathering of data from hospitalised patients by the Committee for Medical Ethics from the Ghent University Hospital (reference number BC-07507) and authorisation for possible individual data linkage using the national register number from the Information Security Committee (ISC) Social Security and Health (reference number IVC/KSZG/20/384). Linkage of hospitalised patient data to vaccination and testing within the LINK-VACC project was approved by the Medical Ethics Committee UZ Brussels VUB on 3 February 2021 (reference number 2020/523), and authorisation from the ISC Social Security and Health (reference number IVC/KSZG/21/034). Denmark: Only administrative register data was used for the study. According to Danish law, ethics approval is exempt for such research, and the Danish Data Protection Agency, which is dedicated ethics and legal oversight body, thus waives ethical approval for the study of administrative register data when no individual contact of participants is necessary, and only aggregate results are included as findings. The study is, therefore, fully compliant with all legal and ethical requirements, and there are no further processes available regarding such studies. Navarre (Spain): The study was approved by Navarres Ethical Committee for Clinical Research, which waived the requirement of obtaining informed consent. Portugal: The study received approval from the Ethical Committee and the Data Protection Officer of the Instituto Nacional de Saude Doutor Ricardo Jorge. Given that data was irreversibly anonymised, the need for the participants informed consent was waived by the Ethical Committee. Italy: This study, based on routinely collected data, will not be submitted for approval to an ethical committee because the dissemination of COVID-19 surveillance data was authorised by the Italian law N. 52 of 19 May 2022, following the law decree N. 24 of 24 March 2022 (Article n. 13). Based on the same acts, the information on COVID-19 vaccination was retrieved by the Italian National Institute of Health using data from the National Immunisation Information System of the Italian Ministry of Health. Because of the retrospective design and the large size of the population under study, in accordance with the Authorisation n. 9 released by the Italian data protection authority on 15 December 2016, the individual informed consent was not requested for the conduction of this study. Sweden: The Swedish study is approved by the Swedish Ethical Review Authority (2020 06859, 2021 02186) and has conformed to the principles embodied in the Declaration of Helsinki. Consent to participate is not applicable as this is a register-based study.

